# Predicting bladder cancer molecular subtypes linked to bacillus Calmette-Guerin response from histology images using deep learning

**DOI:** 10.64898/2026.05.05.26352375

**Authors:** Farbod Khoraminia, Mitchell Olislagers, Florus Christiaan de Jong, Farhan Akram, Alberto Nakauma Gonzalez, Danique Lichtenberg, Andrew Stubbs, James C. Costello, Lucia Rijstenberg, Geert J. L. H. van Leenders, Alina Vrieling, Katja K. H. Aben, Lambertus A. L. M. Kiemeney, Robert F. Hoedemaeker, Chris H. Bangma, Sita Vermeulen, Geert Litjens, Nadieh Khalili, Tahlita C.M. Zuiverloon

## Abstract

**Background and objective:** High⍰risk non⍰muscle⍰invasive bladder cancer (HR⍰NMIBC) is treated with transurethral resection and intravesical BCG instillations, yet ≈50% recur and 20% progress to invasive disease. Although molecular subtyping, e.g., BCG-response-subtype (BRS), is associated with progression risk and may aid risk stratification, yet is costly and time-consuming. Intratumoral heterogeneity complicates accurate subtyping. To address these challenges, we developed a deep-learning model that predicts BRS from routine hematoxylin-eosin-stained images. We verified the model’s area-by-area predictions against tissue-level gene-expression maps.

**Methods and participants:** Hematoxylin-eosin-stained images from 231 HR-NMIBC patients with known BRS were used to develop a deep-learning model through cross-validation, then validated in 83 independent samples. The model’s spatial predictions were assessed using spatial transcriptomics to map gene expression to tissue locations in five HR-NMIBC tumors.

**Outcome measurements and statistical analysis:** Discriminative ability for BRS3 vs. BRS1/2 was measured by AUC. Spatial alignment was assessed by calculating Pearson and Spearman correlation coefficients between model predictions and BRS fractions; significance was assessed through permutation analysis.

**Key findings and limitations:** The trained algorithm achieved AUC of 0.79 (development) and 0.71 (external) to detect BRS3 vs BRS1/2. Tile-level correlation between model output and molecular labels was significant (Pearson r = 0.33–0.44; p ≤ 0.002). Limitations include retrospective sampling and limited spatial transcriptomic cases.

**Conclusions and clinical implications:** Our trained algorithm showed potential to stratify HR⍰NMIBC patients by clinically relevant BCG⍰response subtypes using routine hematoxylin-eosin-stained images and showed predicted spatial heterogeneity comparable to molecular profiling. Prospective validation is required before any clinical implementation.

**Patient summary:** Standard pathology images contain hidden details related to tumor’s molecular subtype. We trained an AI model to read these routine images and identify specific bladder cancer subtypes associated with poor response to BCG therapy. This approach may help reveal molecular subtype-associated information from routine pathology images, without additional laboratory procedures.

## Introduction

Bladder cancers classified as high-risk non-muscle-invasive (HR-NMIBC) carry a high recurrence and progression risk. Standard management is transurethral tumor resection followed by intravesical bacillus Calmette-Guérin (BCG) instillation [1]. BCG is a costly and difficult-to-produce attenuated vaccine, with periodic global shortage, each correlating with higher recurrence rates of bladder tumors [2,3]. Approximately 20% of HR-NMIBC tumors progress to muscle-invasive or metastatic disease, with five-year mortality reaching 50–60% [4]. For BCG-unresponsive HR-NMIBC, guidelines recommend bladder removal, a high-morbidity procedure that reduces quality of life [1,5]. Current risk stratification, based on EORTC tables from 2,596 patients in seven trials with minimal BCG exposure (171 BCG-treated) and on EAU prognostic groups developed from datasets excluding BCG-treated patients, is inadequate to identify patients at the highest BCG failure risk [6]. Consequently, BCG is applied as a one-size-fits-all treatment, causing exposure to unnecessary toxicity and delaying alternative treatment [7]. Improved stratification supports earlier cystectomy when indicated and, once validated, bladder-sparing options [8,9].

Previously, de Jong et al. identified and validated three clinically relevant transcriptomic-based HR-NMIBC subtypes (BCG Response Subtype: BRS1/2/3), with aggressive BRS3 carrying the highest progression risk [10]. While molecular subtyping outperforms clinicopathological features for risk assessment, next-generation sequencing is time-consuming, costly, and needs specialized equipment, hindering clinical implementation [11]. Moreover, bulk sequencing can be confounded by intratumoral heterogeneity, potentially missing focal, therapy-resistant areas [12,13]. These limitations motivate developing more efficient and faster methods that can spatially locate subtypes to refine bulk subtyping and reduce sampling error. Such spatial maps may help flag regions for targeted molecular confirmation. Advances in deep-learning (DL) and digital pathology allow routine hematoxylin-eosin-stained (H&E) slides to yield molecular insights, including microsatellite instability and molecular subtypes, potentially reducing reliance on labor-intensive transcriptomics [14-17]. In NMIBC, prior computational histology AI approaches have shown that H&E-based models can support risk stratification and predict poor BCG response in high-grade Ta disease [18]. Predicting molecular subtype from H&E provides a complementary approach by linking AI predictions to an established biological framework, improving interpretability, enabling spatial assessment of intratumoral heterogeneity, and potentially reducing sequencing costs. In contrast, direct outcome-prediction models may be more clinically actionable but are more vulnerable to treatment patterns, cohort-specific confounding, and endpoint definitions. Thus, both strategies are complementary rather than competing. While DL-based molecular subtyping from H&E has shown promise in other cancers, to our knowledge, its application to predicting molecular subtypes in HR-NMIBC has not been evaluated.

Here, we developed the Bladder Molecular Subtyping Network (BMolNet), a DL model to predict BRS from H&E images in HR-NMIBC patients (Error! Reference source not found.**A, B**). We validated BMolNet through cross-validation and external-validation (**Figure 1B, C**). Additionally, we assessed whether BMolNet can capture intratumor heterogeneity, by generating spatial subtype maps and compared them against spatial transcriptomics-derived labels (Error! Reference source not found.**D**).

**Figure 1:**
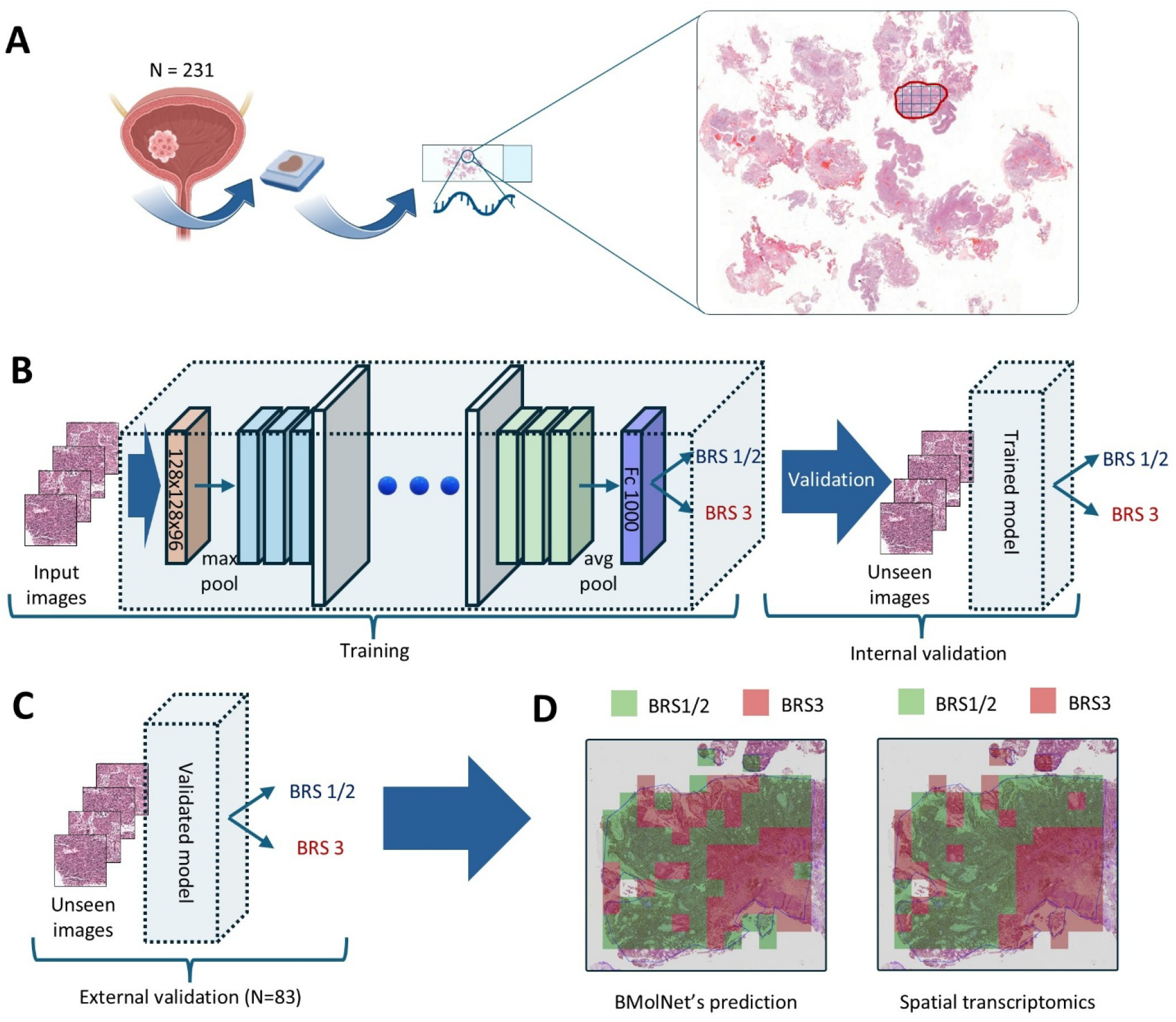
Schematic representation of BMolNet workflow to predict BCG Response Subtype (BRS) in high-risk non-muscle-invasive bladder cancer (HR-NMIBC) patients from hematoxylineosin-stained (H&E)-stained images. (A) Overview of the study cohort: 231 HR-NMIBC patients were enrolled; their H&E-stained tissue sections were digitized, and regions selected for RNA extraction were manually annotated. From these annotations, image tiles were cropped for model development. (B) The cropped image tiles were input into BMolNet, a DL model based on the ConvNeXt architecture. During the training phase, the model learned to classify the images into BCG response subtypes (BRS 1/2 and BRS 3). Internal validation was performed on held-out tiles. (C) External validation workflow: the trained BMolNet model was applied to an independent set of unseen H&E tiles from 83 HR-NMIBC patients. (D) Comparison of BMolNet’s spatial predictions (left) against ground-truth spatial transcriptomics (right), demonstrating concordance and highlighting intratumoral heterogeneity between BRS subtypes (green = BRS 1/2; red = BRS 3).

## Patients and methods

Figure 1 summarizes study design, including cohort selection, model development, external validation, and spatial transcriptomics comparison.

### Patient cohorts

Two independent HR-NMIBC patient cohorts were used: development cohort and external-validation cohort (**Figure 1A-C**). Development cohort included 231 primary, treatment-naïve HR-NMIBC patients (Ta/T1HG) treated with ≥5/6 BCG induction instillations (2000-2018), previously profiled by de Jong et al. using whole-transcriptome sequencing across five hospitals (four in the Netherlands; one in Norway). We collected and scanned corresponding diagnostic H&E slides [10]. Slide review for development cohort was performed in de Jong et al.; for external-validation cohort, a pathologist (LR) applied identical review procedure to identify invasive front (T1) and ≥80% tumor-purity regions (Ta/T1) matching sequenced tissue. External-validation cohort included 83 primary Ta/T1 HR-NMIBC patients (2011-2017) treated with ≥5 BCG induction instillations, drawn from UroLife (prospective cohort across 22 Dutch hospitals) and Nijmegen Bladder Cancer Study (NBCS; across seven Dutch hospitals) [19,20]. Tumor material was retrieved via Dutch Nationwide Pathology Databank (Palga) [21]. Clinical, pathology review, and WTS data were obtained from source studies; additionally, scanned diagnostic H&E slides from external-validation cohort were used. In both cohorts, BCG followed Southwest Oncology Group (SWOG) guidelines and follow-up used European Association of Urology (EAU) guidelines. For development cohort, Institutional Review Board approval was granted by Erasmus MC Medical Research Committee (MEC-2018-1097). For external-validation cohort, Committee for Human Research region Arnhem-Nijmegen approved UroLife (CMO 2013-494) and NBCS (CMO 2005-315) studies.

### Scanning and pre-processing

H&E slides from both cohorts were digitized and processed under a unified protocol. Regions of interest matched sequenced regions, and 10× image tiles were extracted from all sequenced regions per case. F.K. reviewed tile quality and excluded low-quality tiles (**Supplementary Note 1.1**). Development cohort tiles underwent Macenko stain normalization, whereas external cohort was intentionally left unnormalized to preserve site-dependent variability [22]. Full scanning, example annotations, tiling, QC, and normalization details are provided in **Supplementary Note 1.1**. and **Supplementary Figure 1**).

### Model development

Because BRS1 and BRS2 show similar clinical behavior, they were grouped (BRS1/2) and contrasted with aggressive BRS3. A ConvNeXt-based classifier was trained on patient-level labels [23], using stratified 5-fold cross-validation to prevent leakage (statistical analysis in **Supplementary Figure 3**). Within folds, we selected the model with lowest validation tile-loss. Tile probabilities were binarized using a Youden index-optimized threshold; patient-level classes were then determined by majority vote and mean tile probability. We evaluated (i) discrimination (ROC-AUC), (ii) correlation with bulk transcriptomic–derived BRS3 fractions, and (iii) alignment with spatial transcriptomics labels (**Supplementary Note 1.2**). Training protocol and hyperparameters are in **Supplementary Note 1.1**.

### Gene expression profiling

#### Tumor heterogeneity measured from whole-transcriptome data

To quantify intratumoral heterogeneity from bulk RNA-seq, we estimated sample-wise BRS1-3 fractions from whole-transcriptome data. Differentially expressed protein-coding BRS-associated genes were identified using DESeq2 (log2 fold change > 1.0; Benjamini–Hochberg adjusted p < 0.01) was performed per batch as defined by de Jong et al., retaining only overlapping genes [10,24]. Raw counts were cohort-normalized using DESeq2 variance-stabilizing transformation, median-centered, and gene-set means were converted to proportional fractions (set to zero when group mean < −1.0). Concordance was defined as agreement between majority-fraction subtype and the bulk-assigned subtype. Progression-free survival (PFS; time from diagnosis to progression to ≥T2, lymph node or distant metastasis) and overall survival (OS; time from diagnosis to death from any cause) were evaluated with Kaplan– Meier curves and two-sided log-rank tests; stratifying “high/low” by the median BRS3 fraction within BRS1 samples and censoring at last follow-up (**Supplementary Note 1.3**).

#### Spatial transcriptomics

Five external-cohort samples underwent Visium 10X spatial transcriptomics with CystAssist spot-level BRS labels (**Figure 1D; Supplementary Note 1.3**). For comparison with BMolNet, matching H&E sections were scanned with Axioscan, converted from CZI to TIFF using our public converter (https://github.com/farbodkhoraminia/CZI-converter), and tiled at 10× into non-overlapping 512 × 512 px tiles. Tile BRS classes were assigned by majority vote across overlapping spots, and tile-level BRS3 fraction by averaging spot labels. These tile-level labels/fractions served as spatial reference for BMolNet output. Procedural details, including sequencing protocols and QC thresholds, are in **Supplementary Note 1.3**.

#### Statistical analysis

Discrimination was assessed using ROC–AUC at tile and patient levels (cross-validation and external-validation). Spatial concordance methods and significance testing are described in **Supplementary Note 1.2**.

## Results

### BMolNet predicts BRS molecular subtypes from histology images

A CONSORT chart for HR-NMIBC patients used in training and validation is shown in **Supplementary Figure 2**. Of 283 patients with known molecular subtypes, 21 slides were unscannable because of physical damage, 31 slides were blurry, leaving 231 patients for analysis (**Table 1**). **Error! Reference source not found**.Cohort composition by BRS1/2 versus BRS3 is summarized in **Table 2**. Across folds, 71–74% of patients were BRS1/2; the external cohort included 58 BRS1/2 (70%) and 25 BRS3 (30%). Training sets contained 4,041–4,638 tiles (mean 4,368; SD 219). Internal validation sets included 845–1,399 BRS1/2 and 272–514 BRS3 tiles; the external set had 2,155 BRS1/2 and 1,566 BRS3 tiles. Tiles per patient did not differ across folds (p=0.41; **Supplementary Figure 3**).

**Table 1:** Clinical overview of development cohort and external-validation cohort.

| Parameter | Category | Development cohort | External-validation cohort | P value |
| --- | --- | --- | --- | --- |
| Grade | G2 | 13 (5.6%) | 7 (8.4%) | 0.52 |
|  | G3 | 218 (94.4%) | 76 (91.6%) |  |
| Stage | Ta | 36 (15.6%) | 4 (4.8%) | 0.012 |
|  | T1 | 195 (84.4%) | 79 (95.2%) |  |
| Age (years) | Median (IQR) | 70.00 (62.00-77.00) | 68.00 (62.50-72.50) | 0.037 |
| Concomitant CiS | No | 183 (79.2%) | 80 (96.4%) | <0.001 |
|  | Yes | 48 (20.8%) | 3 (3.6%) |  |
| re-TURB | No | 74 (32.0%) | 1 (1.2%) | <0.001 |
|  | Yes | 157 (68.0%) | 82 (98.8%) |  |
| Gender | Male | 185 (80.1%) | 67 (80.7%) | 1.0 |
|  | Female | 46 (19.9%) | 16 (19.3%) |  |
| Tumor size | ≤30 | 39 (16.9%) | 4 (4.8%) | 1.0 |
|  | >30 | 38 (16.5%) | 3 (3.6%) |  |
|  | Missing | 154 (66.7%) | 76 (91.6%) |  |
| Time to end of follow-up (months) | Median (IQR) | 67.00 (33.50-98.00) | 61.76 (47.13-68.05) | 0.011 |
| Progression | No | 150 (64.9%) | 64 (77.1%) | 0.057 |
|  | Yes | 81 (35.1%) | 19 (22.9%) |  |
| Continuous comparisons: Mann–Whitney U test; categorical comparisons: Chi-square or Fisher exact where counts <5; two-sided p values reported. Categories marked “Missing” were excluded from hypothesis tests |  |  |  |  |

**Table 2:** Overview of patients and tiles distribution across folds for BRS1/2 and BRS3 groups.

| Category | Set | BRS group | All unique patients/tiles (%) | Mean ± SD across 5 folds |
| --- | --- | --- | --- | --- |
| Patients | Training | BRS1/2 | 166 (72%) | 132.8 ± 1.1 |
| | | BRS3 | 65 (28%) | 52.0 ± 1.0 |
| | Internal validation | BRS1/2 | 166 (72%) | 33.2 ± 1.1 |
| | | BRS3 | 65 (28%) | 13.0 ± 1.0 |
|  | External-validation | BRS1/2 | 58 (70%) | NA (no folds) |
|  |  | BRS3 | 25 (30%) | NA (no folds) |
| Tiles | Training | BRS1/2 | 5552 (76%) | 4441.6 ± 223.7 |
| | | BRS3 | 1756 (24%) | 1404.8 ± 105.0 |
| | Internal validation | BRS1/2 | 5552 (76%) | 1110.4 ± 223.7 |
| | | BRS3 | 1756 (24%) | 351.2 ± 105.0 |
|  | External-validation | BRS1/2 | 2155 (58%) | NA (no folds) |
|  |  | BRS3 | 1566 (42%) | NA (no folds) |
| BRS = Bacillus Calmette-Guérin Response Subtype; Five-fold cross-validation was used. “Mean ± SD across 5 folds” summarizes the five training-fold counts for each class; for training rows, “All Patients / Tiles (unique)” reflects the distinct individuals or image tiles ($\Sigma$ across folds ÷ 4, because each sample appears in four of the five training folds); internal-validation totals are unique (each sample appears in exactly one fold); external-validation totals are independent of the CV splits and therefore have no fold statistics | | | | |

Across five folds, BMolNet achieved macro-averaged AUC 0.79 (fold AUCs: 0.77, 0.67, 0.74, 0.92, 0.86; **Figure 2A**), and external-validation AUC 0.71 (**Figure 2B**). The pooled cross-validation confusion matrix indicated 136 BRS1/2 and 40 BRS3 correctly classified, with 30 BRS1/2 and 25 BRS3 misclassified; mean balanced-accuracy 0.72 ± 0.09 (**Figure C**). External-validation achieved balanced accuracy 0.63 with 41/58 BRS1/2 and 25/36 BRS3 correctly classified (**Figure 2D**).

**Figure 2:**
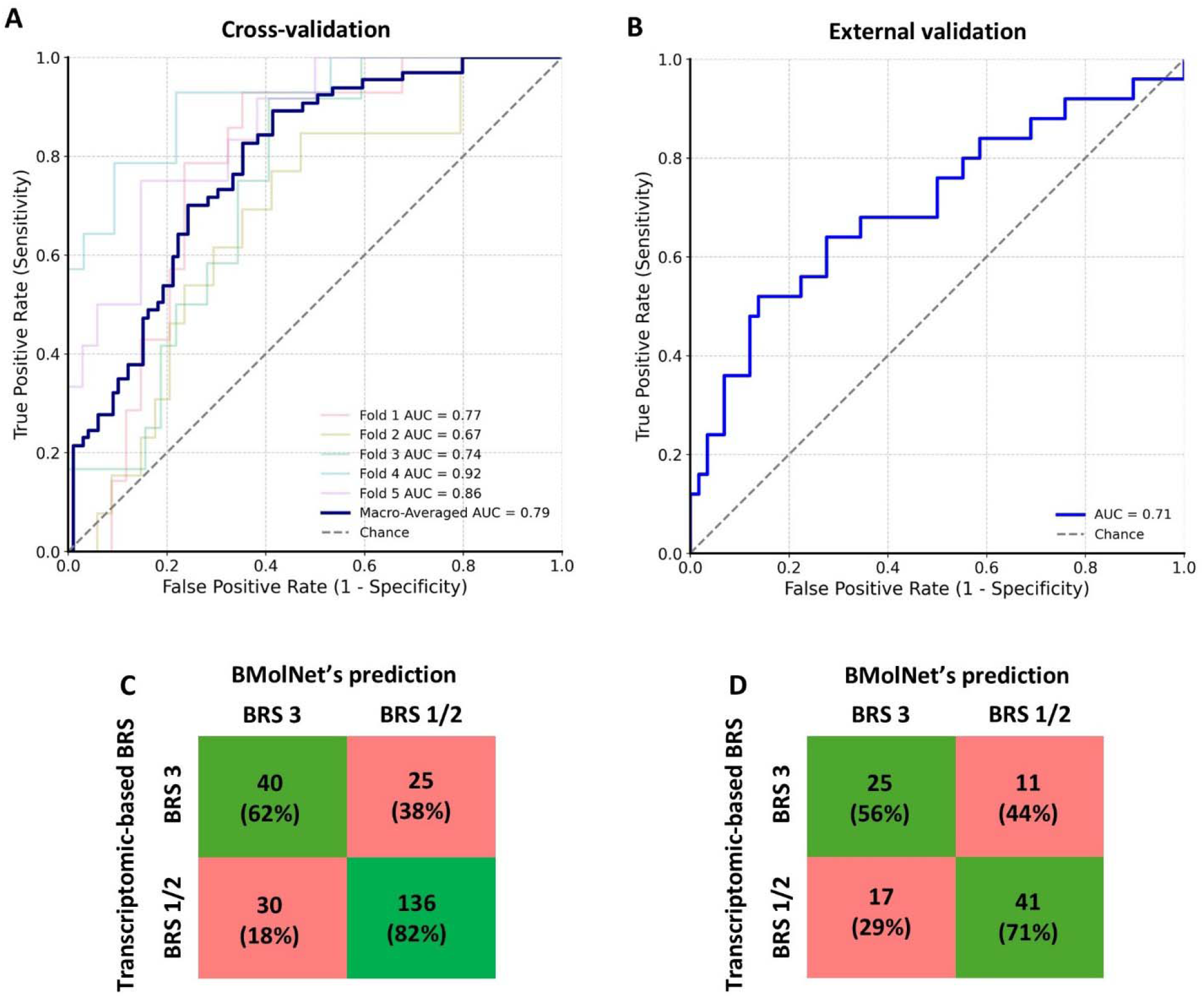
(A) Macro-averaged Receiver–operating characteristic (ROC) curves for the cross⍰/validation cohort, where each curve represents performance on one held⍰/out fold and the thick line shows the macro⍰/averaged AUC. (B) ROC curve for the external-validation cohort, with the overall AUC reported. (C) Cross-validation confusion matrix with transcriptomic BRS labels on the y-axis and BMolNet predictions on the x-axis. Each cell shows the number of patients and the percentage within that ground-truth (row) group; green = correct, red = incorrect. (D) Confusion matrix for the external-validation cohort, showing counts of true versus predicted BRS subtypes.

### BMolNet can predict intratumor heterogeneity

Bladder cancer is known for intratumoral subtype heterogeneity, which may influence prognosis and treatment response [12,25-27]. Hence, we investigated whether multiple subtypes presence within one patient has clinical impact. We estimated heterogeneity and identified a BRS1 subgroup with high BRS3 fraction (upper 50%), which showed worse OS (p = 0.026) and PFS (p = 0.0089) than the lower 50% (**Supplementary Figure 4A**). Since subtype heterogeneity is clinically relevant, we asked whether BMolNet can localize mixed BRS subtypes at tile level by validating predictions against spatial-transcriptomics–derived labels. To our knowledge, this is the first HR-NMIBC study to train a DL model on bulk transcriptomics-defined subtypes and validate its ability to detect intratumor heterogeneity.

Bulk transcriptomics provided heterogeneity reference. Fraction-based BRS was concordant with bulk-assigned labels 87.7% overall (90% for BRS3), and most tumors showed some heterogeneity (**Supplementary Figure 4B**). Error! Reference source not found.To test whether BMolNet predicted probabilities indicate subtype heterogeneity, we correlated patient-level mean predicted BRS3 probability with the transcriptomic-derived BRS3 fraction (**Supplementary Figure 5A, C**). Patient-level mean predicted BRS3 probability correlated with transcriptome-derived BRS3 fraction (out-of-fold CV: Pearson r = 0.44 and Spearman ρ = 0.38; both p < 0.001; **Supplementary Figure 5A, D**). When patients were grouped into ten bins increasing BRS3 fraction, mean predicted probability rose across bins (**Supplementary Figure 5B, E**). Against 10000 label shuffles, correlations were significantly larger (**Supplementary Figure 5C, F; permutation p < 1×10^−4^**). For spatial validation, we co-registered Visium spots to WSIs and compared tile-level BRS3 fractions with BMolNet probabilities (**Supplementary Table 1**). BMolNet heatmaps recapitulated ST patterns (**Figure 3; Supplementary Figure 6**) showing positive concordance (mean Pearson r ≈ 0.33 ± 0.22), with similar probability distributions (mean Jensen–Shannon distance ≈ 0.23 ± 0.04). Alignment was significant in four of five samples (permutation p < 1×10^−4^). Full metrics are in **Supplementary Note 1.2** and **Supplementary Table 1**.

**Figure 3:**
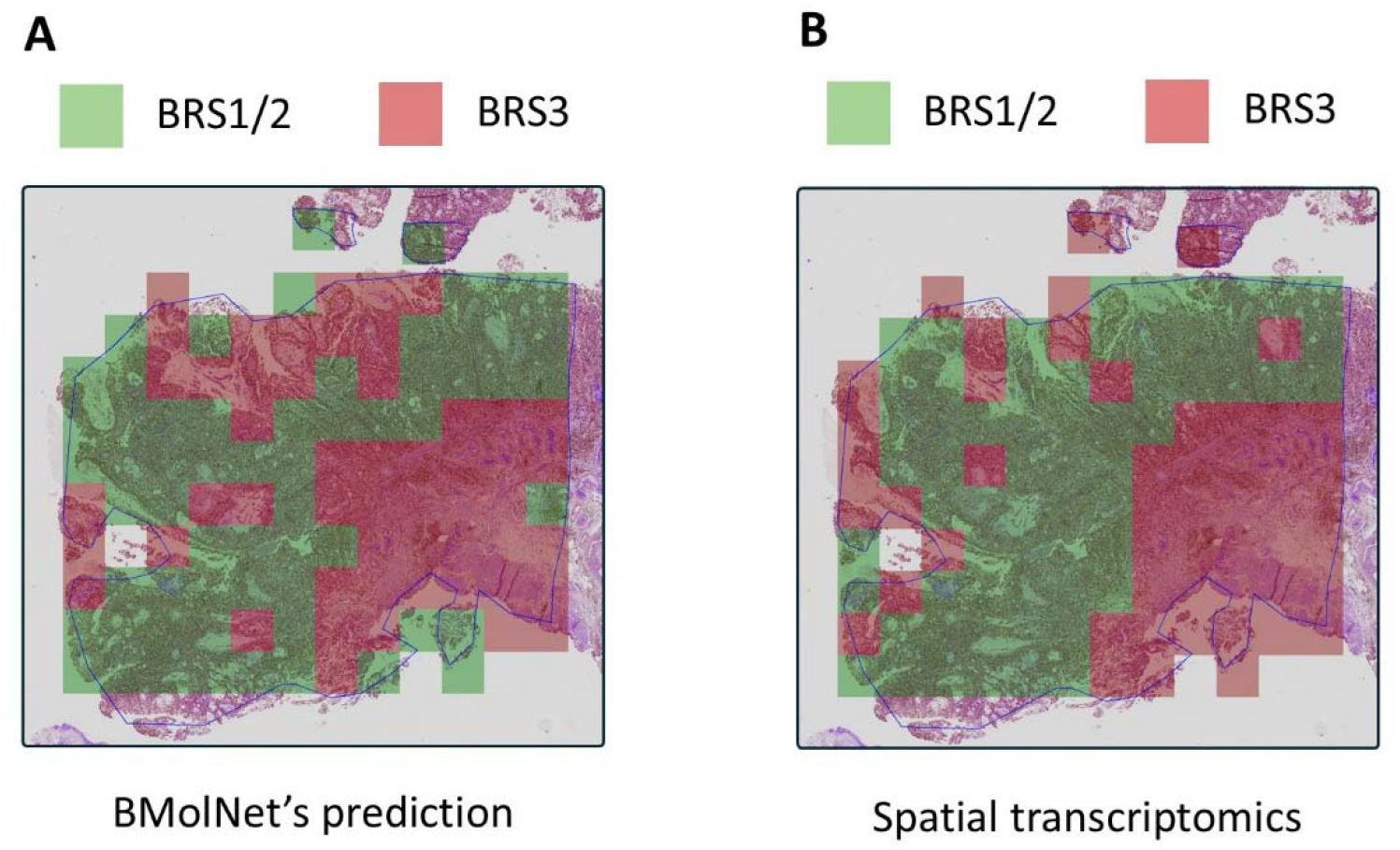
Spatial overlay of BMolNet predictions versus transcriptomic ground truth on an H&E-stained image. (A) MolNet tile-level predictions overlaid on the H&E within the annotated region (blue outline). (B) Corresponding molecular BRS assignment derived from spatial transcriptomics for the same region.

## Discussion

Despite their prognostic value, BRS classes are not routinely available at diagnosis. We developed and externally validated BMolNet, an H&E-based DL model that predicts BRS subtypes and aligns with spatial transcriptomics. Because it adds no wet-lab time, BMolNet can deliver results the same day (≤∼1 h), whereas NGS takes days to weeks. Early BRS3 identification can assist pathologists in prioritizing cases for further review or guide confirmatory assays (p53 IHC, FGFR3 testing, targeted NGS). BMolNet extends prior H&E-based computational pathology work in NMIBC by focusing on transcriptomically defined BCG-response subtypes rather than direct clinical outcome prediction, and by evaluating spatial correspondence with spatial transcriptomics [18]. This biological anchoring may improve interpretability and enable spatial mapping of subtype-associated intratumoral heterogeneity. Direct outcome-prediction models may be more closely aligned with treatment decisions but can be more sensitive to treatment patterns, endpoint definitions, and cohort-specific confounding. These strategies are complementary. This study should be interpreted as proof-of-concept and hypothesis-generating, not as evidence for treatment-guiding implementation.

BMolNet achieved AUC 0.79 in 5-fold cross-validation (balanced-accuracy 0.72). In external cohort, performance was similar (AUC0.71; balanced-accuracy 0.63), with BRS3 sensitivity 0.69 (25/36) and BRS1/2 specificity 0.71 (41/58). External-validation used slides from different hospitals and years and intentionally left them unnormalized to mirror routine practice. The ∼0.05–0.10 AUC reduction likely reflects inter-center stains and scanners variability, a pattern also reported in deployed computational pathology systems. This performance supports model’s robustness, further prospective, multi-center evaluation across diverse patient populations. A potential future use of BMolNet is as a triage tool rather than a standalone clinical decision system. Because it runs on routinely acquired H&E slides, it could provide subtype probabilities and heatmaps in ∼1 hour, bypassing heavy logistics and transcriptomics or multiplex immunofluorescence delay. In a potential workflow, pathologists could use heatmaps to identify BRS3-predominant regions for confirmatory assessment (e.g., p53 IHC, FGFR3 testing, targeted RNA profiling), interpreted alongside clinicopathological, imaging, and molecular findings in multidisciplinary review. Thresholds could be adapted to the intended use case, for example, by prioritizing sensitivity for confirmatory testing. However, clinical utility, safety, and generalizability require prospective validation. This spatial use case is particularly relevant because bladder cancer shows intratumoral heterogeneity, which bulk subtyping may miss and which can increase sampling error in risk assessment [12,25,28]. Explainability tools, including class activation maps or attention overlays, may help highlight candidate regions for further molecular or pathological investigation [29-31]. In our analysis, tumors with higher BRS3 fraction had worse outcomes even when bulk transcriptomics classified them as BRS1, whereas this pattern was not evident within bulk BRS2. Although mechanisms were not evaluated, one possible explanation is that BRS2 and BRS3 share progression-linked pathways, but BRS2 may remain partly controlled by immune activity, while BRS3 is associated with an immunosuppressive microenvironment [10]. Conversely, BRS1 is biologically more distinct from BRS3, which may explain the stronger prognostic impact of BRS3 admixture within BRS1.

BMolNet’s patient-level probabilities correlated with BRS fraction (ross-validation: Pearson r = 0.44, Spearman ρ = 0.38; external: Pearson r = 0.33; Spearman ρ = 0.32; all p ≤ 0.01). This indicates that BMolNet captures bulk-derived BRS3 signal and may help identify cases warranting further biological characterization or confirmatory assessment in future validation studies. Additionally, BMolNet’s spatial overlay aligns with spatial RNA maps in both homogeneous and mixed regions that mark intratumor heterogeneity (**Figure 3; Supplementary Figure 6**), quantitatively with significant agreement in most cases (low JSD and comparable entropy shown in **Supplementary Note 1.2; Supplementary Table 1**). Although Visium set was small (n = 5), this provides orthogonal support for using heatmaps to target confirmatory assays. Our small-scale exploratory Visium analysis suggests BRS signature is detectable at spot resolution, with BRS3 signals appearing in CAF-rich stroma (**Supplementary Note 1.3**). Future studies can pair heatmaps with targeted RNA/IHC panels, such as CAF markers, to confirm spatial co-localization and prognostic value. These findings suggest BMolNet can both predict patient-level BRS3, and locate BRS3 regions. This spatial anchoring could address a bulk subtyping limitation—sampling bias—by directing which tissue region to sequence.

Limitations include the retrospective design, moderate external performance, and the small spatial validation set. Because the model was trained on pathologist-selected high-tumor-content regions, performance on whole-slide scans, biopsies, artifact-rich tissue, and more heterogeneous real-world specimens remain unknown. Thus, present findings are proof-of-concept and require prospective validation and clinical utility assessment before implementation. Future work should link BMolNet heatmaps to histologic features and molecular markers, such as nuclear atypia, immune infiltrates, Ki-67, PD-L1, or CAF markers, to improve biological interpretability and clinical trust. Multi-regional sampling and orthogonal spatial assays should test whether BMolNet consistently localizes high-risk BRS3 tissue within heterogeneous tumors. Pathology-specific foundation model initialization may further improve robustness and generalizability [32-34]. Finally, BMolNet offers a ready foundation to transfer model’s learned features and fine-tune on other HR-NMIBC cohorts to predict clinically relevant molecular subtypes from H&E images.

## Conclusion

BMolNet turns routine slides into subtype probabilities and region-level signals. With broader, prospective cohorts—and stronger backbones from pathology foundation models—the same pipeline may extend across taxonomies and settings. The promise is a single, low-cost entry point to molecular risk and intratumor heterogeneity. Now it needs clinical proof.

## Data Availability

The histopathology images and molecular data generated for this study are available from the corresponding author on reasonable request. These data were also used in the CHIMERA Grand Challenge (Task 2). Copies of CHIMERA training files are hosted via the CHIMERA portal and the AWS Open Data Registry under the CHIMERA terms. The BMolNet code will be made available upon manuscript acceptance at https://github.com/erasmus-ur/BMolNet.

## Data and code availability

Acknowledgments

This work was funded by the EU Horizon 2020 Marie Skłodowska-Curie grant No. 860627 (CLARIFY). The authors express their gratitude to Emiel Janssen, Vebjørn Kvikstad, Jolien Mensink, Sébastien Rinaldetti for their help with data collection and data handling. We thank the Erasmus MC Pathology Research and Trial Service (PARTS) team—Angelique van der Made, Thierry P.P. van den Bosch, Karishma A. Lila, and Marit de Haan—for their support with sample handling and data acquisition. We also acknowledge Eric Bindels’ group, and in particular Gregory van Beek and Mathijs Sanders, for their assistance with data acquisition and technical support. We thank other members of the Erasmus Urothelial Cancer Research Group, and Radboud UMC Computational Pathology Group, especially Tokameh Mahmoudi, Vera Rutten, Joost Boormans, Shahla Romal, Mathijs Scholtes, and Stephan Dooper, for their constructive ideas. SHV is supported by a grant from the Netherlands Organization for Scientific Research (NWO Vidi 91717334). The UroLife study was financially supported by Alpe d’HuZes/Dutch Cancer Society (KUN 2013-5926) and Dutch Cancer Society (2017-2/11179). The authors thank all the patients who participated in UroLife and thank the following hospitals for their involvement in recruitment for the UroLife study: Amphia Ziekenhuis, Breda/Oosterhout (D.K.E. van der Schoot); Ziekenhuis Bernhoven, Uden (A.Q.H.J. Niemer); Canisius-Wilhelmina Ziekenhuis, Nijmegen (D.M. Somford); Catharina Ziekenhuis, Eindhoven (E.L. Koldewijn); Deventer Ziekenhuis, Deventer (P.L.M. van den Tillaar); Elkerliek Ziekenhuis, Helmond (E.W. Stapper, P.J. van Hest); Gelre Ziekenhuizen, Apeldoorn/Zutphen (D.M. Bochove-Overgaauw); Isala Klinieken, Zwolle (E. te Slaa); Jeroen Bosch Ziekenhuis, ‘s-Hertogenbosch (S. van der Meer); Meander Medisch Centrum, Amersfoort (F.S. van Rey); Medisch Spectrum Twente, Enschede (M. Asselman); Maxima Medisch Centrum, Veldhoven/Eindhoven (L.M.C.L. Fossion, K. de Laet); Maasziekenhuis Pantein, Boxmeer (E. van Boven); Radboudumc, Nijmegen; Rijnstate, Arnhem/Velp/Zevenaar (C.J. Wijburg); Slingeland Ziekenhuis, Doetinchem (A.D.H. Geboers); St. Anna Ziekenhuis, Geldrop (A. Sonneveld); Elisabeth-TweeSteden Ziekenhuis, Tilburg/Waalwijk (P.J.M. Kil, B.P. Wijsman); St. Jansdal Ziekenhuis, Harderwijk (W.J. Kniestedt); VieCuri, Venlo (G. Yurdakul, A.H.P. Meier); Ziekenhuis Gelderse Vallei, Ede (M.D.H. Kortleve); Ziekenhuisgroep Twente, Almelo/Hengelo (E.B. Cornel). The authors thank the registration team of the Netherlands Comprehensive Cancer Organisation (IKNL) for the collection of data for the Netherlands Cancer Registry as well as IKNL staff for scientific advice.

## Declaration

During preparation, the first author used ChatGPT to improve readability, then reviewed and edited the text and takes full responsibility for the content.

## Notes

### Competing Interest Statement

The authors have declared no competing interest.

### Funding Statement

This work was funded by the EU Horizon 2020 Marie Skłodowska-Curie grant No. 860627 (CLARIFY). SHV is supported by a grant from the Netherlands Organization for Scientific Research (NWO Vidi 91717334). The UroLife study was financially supported by Alpe d HuZes/Dutch Cancer Society (KUN 2013-5926) and Dutch Cancer Society (2017-2/11179).

### Author Declarations

Erasmus MC Medical Research Committee of Erasmus MC gave ethical approval for this work (MEC-2018-1097). Committee for Human Research region Arnhem-Nijmegen gave ethical approval for the UroLife study (CMO 2013-494) and the Nijmegen Bladder Cancer Study (CMO 2005-315).

