## Supplementary Table 1 for "Predicting bladder cancer molecular subtypes linked to bacillus Calmette-Guerin response from histology images using deep learning"

**Supplementary Table 1: Summary of spatial transcriptomic alignment metrics**

| *Supplementary Table 1. Summary of spatial transcriptomic alignment metrics. The table summarizes eight metrics for five spatial transcriptomic samples. The Jensen–Shannon distance quantifies divergence between BMolNet predictions and BRS fractions. Entropy values reflect complexity, and correlation coefficients (Pearson & Spearman) assess tile-wise agreement under real and randomized conditions.* | | | | | | | | |
| --- | --- | --- | --- | --- | --- | --- | --- | --- |
| All Tumor Sample ID | JSD | Entropy BMolNet | Entropy BRS fraction | Entropy BMolNet / BRS fraction | Pearson correlation actual | Pearson correlation permutation | Spearman correlation actual | Spearman correlation permutation |
| Sample 1 | 0,21 | 2,81 | 3,69 | 0,76 | 0,74 | 0 | 0,70 | 0,02 |
| Sample 2 | 0,25 | 3,71 | 3,69 | 1,01 | 0,13 | 0,08 | 0,05 | 0,04 |
| Sample 3 | 0,29 | 2,31 | 3,76 | 0,62 | 0,40 | -0,11 | 0,36 | -0,06 |
| Sample 4 | 0,18 | 3,29 | 2,62 | 1,25 | 0,17 | -0,16 | 0,12 | -0,11 |
| Sample 5 | 0,20 | 3,32 | 2,71 | 1,22 | 0,21 | 0 | 0,17 | -0,05 |
| Average  (std) | 0,23 (0,04) | 3,09 (0,48) | 3,29 (0,52) | 0,97 (0,25) | 0,33 (0,22) | -0,04 (0,11) | 0,28 (0,23) | -0,03 (0,06) |
| JSD: Jansen-Shannon distance between BRS fraction and BMolNet predicted probability; std: standard deviation | | | | | | | | |
