## Supplementary Note 1 for "Predicting bladder cancer molecular subtypes linked to bacillus Calmette-Guerin response from histology images using deep learning"

**Supplementary Note 1.1: Scanning, pre-processing, and model development methods.**

All H&E slides were digitized on a 3DHistech P1000 and blurry whole-slide images were excluded after visual review. In the development cohort, ROIs corresponding to whole-transcriptome sequencing samples were pixel-wise annotated in QuPath [9]; the external cohort was processed and annotated using the same protocol. Annotations were performed by FK and DL and validated by an expert uropathologist (Prof. Van Leenders). From annotated ROIs, 512×512-pixel tiles were extracted at 10× magnification with 25% overlap; tiles with <50% annotated mask were discarded. Development-cohort tiles underwent Macenko stain normalization using a single reference tile selected by a uropathologist from an independent cohort [21]; no retrospective stain normalization was applied to the external cohort. Tiles showing excessive blood, cautery artifacts, or blurriness were removed after visual review by FK. QuPath was used for annotation and the downstream tiling, QC and normalization steps followed our previously described pipeline [9].

We used a ConvNeXt backbone [22] initialized from ImageNet with earlier blocks frozen and the last 10 blocks of the final stage trainable; a custom classification head with batch normalization was attached. Within each patient-stratified CV fold, tiles were split 80/20 by patient for training/validation. Training augmentation consisted of random horizontal and/or vertical flips; all originals and augmented tiles were used for training, while internal validation and the external test sets were not augmented. To address class imbalance, the majority class (BRS1/2) was randomly duplicated by 15% and the minority class (BRS3) was oversampled to achieve approximate balance in the training set. Optimization used the Adam optimizer with binary cross-entropy-with-logits loss, mixed-precision training, gradient clipping (max-norm 1) and early stopping with 15-epoch patience; weight decay (applied via the optimizer’s weight_decay) was tuned between 1e-5 and 1e-1 (no additional explicit L2 term in the loss). Hyperparameters were selected using a Bayesian search (30 trials per fold) exploring learning rates (1e-4, 1e-3, 1e-2), batch normalization on/off, batch sizes 64–256 (step 64), weight decay 1e-5–1e-1, and scheduler gamma 0.1–0.9 (step 0.1); the best checkpoint per fold was chosen by lowest validation patch-level loss. Tile probabilities were binarized using a Youden-index–optimised threshold on the validation cohort; patient-level class was assigned by majority vote across binarized tiles and patient-level probability was computed as the mean of tile probabilities. All experiments were implemented in Python 3.8 with PyTorch and run on desktop workstations (NVIDIA RTX 4090, Intel® Core™ i9-13900K). Source code is available at https://github.com/farbodkhoraminia/BMolNet

.**Supplementary Note 1.2: Spatial concordance of BMolNet with Visium transcriptomics**

This note provides the full technical description and results for the spatial concordance analysis comparing BMolNet’s tile-level BRS3 probabilities with spatial transcriptomic (Visium) estimates of BRS3 at matched locations. The aim was to quantify how closely model-derived probability maps reflect spatial variation in molecular subtype, beyond slide-level classification. Five tumours from the external-validation cohort underwent Visium CytAssist spot-level profiling with matched H&E sections from the same regions of interest. H&E whole-slide images were scanned on a Zeiss Axioscan (CZI format) and converted to TIFF using our public converter (<https://github.com/farbodkhoraminia/CZI-converter>). Regions of interest were annotated in ASAP (https://github.com/computationalpathologygroup/ASAP). We extracted non-overlapping tiles at 10× magnification (512 × 512 px) using the pathology-whole-slide-data package (<https://github.com/DIAGNijmegen/pathology-whole-slide-data>). Visium spot locations were manually co-registered to the H&E image with a napari-based procedure to ensure spatial correspondence between spots and tiles.

For each H&E tile, we derived a Visium-based reference by averaging the BRS3 labels of all overlapping spots to obtain a tile-level BRS3 fraction and, in parallel, assigning a hard BRS class by majority vote. These fractions and classes formed the spatial ground truth for comparison with BMolNet’s tile-level BRS3 probabilities. Spatial agreement was quantified in three complementary ways: first, tile-wise association, calculated per tumour using Pearson and Spearman correlations between BMolNet probabilities and Visium-derived fractions; second, distributional similarity, assessed by the Jensen–Shannon distance (JSD) between the two spatial distributions, with lower JSD indicating closer alignment; and third, map complexity, evaluated by constructing histograms (bin width 0.05) of each measure, computing Shannon entropy for both, and reporting the entropy ratio (BMolNet/Visium) to summarise relative spatial complexity.To determine whether observed tile-wise correlations exceeded chance, we generated null distributions via 10 000 permutations for each tumour by randomly shuffling BMolNet tile-level probabilities while holding Visium fractions fixed. We then compared true vs. permuted correlations using two-sided Wilcoxon tests. We also report the permutation means ± SD for both Pearson and Spearman.

Spatial trsanscriptomics and corresponding BMOlNet prediction overlays for all five tumors are shown in **Figure 3** and **Supplementary Figure 6**). Tile-wise correlations under true labels averaged Pearson 0.33 ± 0.22 and Spearman 0.28 ± 0.23 across the five tumours, compared with permutation baselines of −0.04 ± 0.11 (Pearson) and −0.03 ± 0.06 (Spearman). JSD which shows divergence between two spatial distribution ranged 0.18–0.29 (mean 0.23 ± 0.04). Entropy which shows complexity 3.09 ± 0.48 for the BMolNet prediction, and was 3.29 ± 0.52 for the Visium labels, resulting in entropy ratio of 0.97 ± 0.25. Four out of five tumors showed correlations significantly greater than chance (p < 1 × 10⁻⁴), while 1/5 did not reach significance (p = 0.45). We provide the full per-sample metrics, JSD, entropy for both maps, entropy ratio, Pearson/Spearman (true) with corresponding permutation means ± SD, summarised in **Supplementary Table 1**, with the cohort-level mean ± SD in the final row.

**Supplementary Note 1.3: Gene expression profiling, heterogeneity estimation, and spatial transcriptomics analysis**

To quantify within-sample heterogeneity from bulk RNA-seq and to provide a reference for model evaluation, we estimated per-sample fractions of BRS1, BRS2 and BRS3 by first identifying differentially expressed genes associated with each BRS group. For this, all tumor samples assigned to a given BRS group were compared against samples from the other two groups using DESeq2; all protein-coding genes from the whole-genome transcriptomics were analysed and differentially expressed genes were defined as those with log2 fold change > 1.0 and Benjamini–Hochberg adjusted p < 0.01 [23]. To avoid batch effects from differences in sequencing workflows, this differential-expression step was done separately per subset of samples (cohort A and B) as defined by de Jong et al., and only genes that overlapped across the subsets were selected as BRS-associated [9] . Raw counts were normalized per cohort following the de Jong procedure using DESeq2 with variance-stabilizing transformation, retaining only protein-coding genes for downstream analysis; normalized gene expression was median-cantered. For each sample we computed a raw score per BRS group as the mean of its marker-gene expression (variance-stabilized, median-cantered). Raw scores < 0 were truncated to 0 and then normalized so that the three BRS scores summed to 1, yielding percentage fractions (0–100%). For visualization (Fig. S3), when truncation caused the three fractions to sum < 100%, the residual was displayed as “other/undetermined”; all statistical analyses used only the BRS1/2/3 fractions. Progression-free survival (PFS) was defined as time from diagnosis to progression to muscle-invasive disease (≥T2), lymph node or distant metastasis; overall survival (OS) was defined as time from diagnosis to death from any cause. Survival analysis was performed using the Kaplan–Meier method and two-sided log-rank tests to compare survival distributions. Patients were stratified into “high” and “low” groups based on the median fraction of the high-risk BRS3 subtype within BRS1 samples; Kaplan–Meier curves for PFS and OS were generated with censoring at last follow-up and compared by two-sided log-rank tests (significance threshold p < 0.05). Analyses were implemented in Python using lifelines v0.26.4 and Matplotlib v3.4.3.

We analyzed spatial gene expression in five treatment-naïve HR-NMIBC FFPE tissue specimens selected by a uropathologist for invasive papillary tumour regions, using the 10x Genomics Visium CytAssist platform. Visium spot libraries were processed and normalized in Seurat and clustered following standard workflows; because not every signature gene is expressed in every spot, we applied the published BRS classifier using only the BRS signature genes that were detected in each individual spot. To benchmark this spot-wise approach, we re-predicted BRS labels for the 151 bulk RNA-seq HR-NMIBC samples using the same spot-specific gene subsets and compared those predictions with the known bulk BRS assignments. Tumour purity per spot was estimated with ESTIMATE, and spots with tumour purity < 0.88 were considered to contain non-tumour cells and treated accordingly in downstream analyses.

In total, 12,472 spots were analyzed. Concordance between known BRSs and the re-classified BRSs using the expressed genes from each of the 12,472 spots had a mean of 84%, with SD 2.9 (total classifications: 12,472*151=1,883,272). Spots classified as BRS3 were significantly enriched in a cluster characterized by overexpression of CAF marker genes (p<0.001). However, when spots with low tumor purity were excluded, spots composed of BRS3 tumor cells were found to be in close spatial proximity to stromal regions.
