## Supplementary Figure 1 for "Predicting bladder cancer molecular subtypes linked to bacillus Calmette-Guerin response from histology images using deep learning"

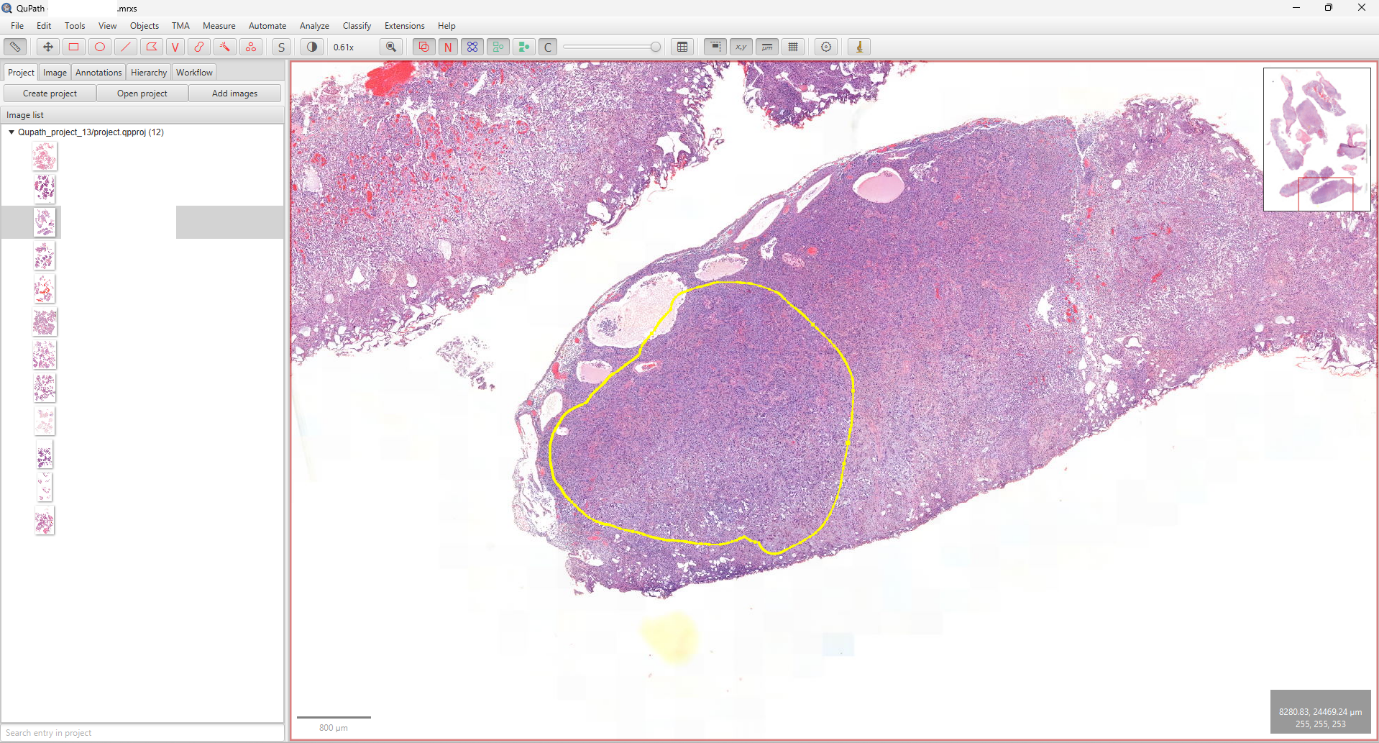
**A**

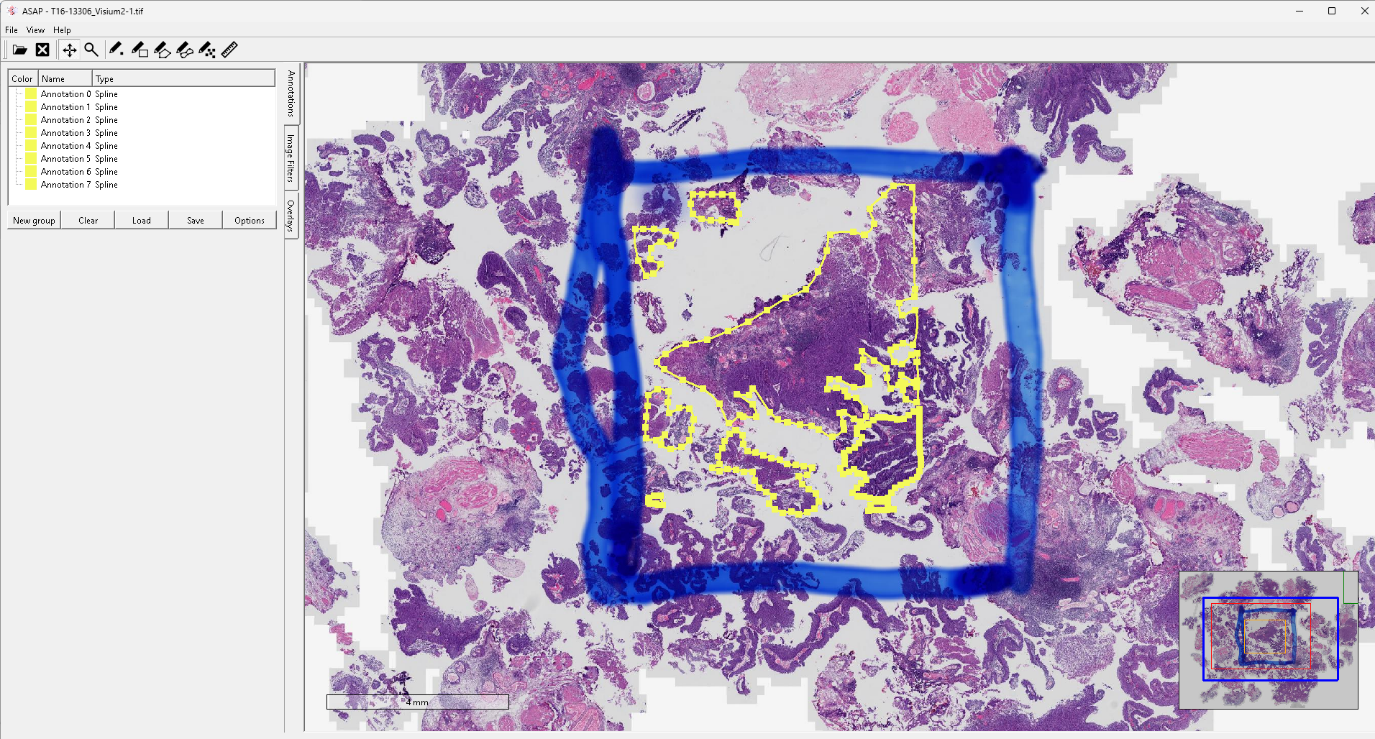
**B**

Supplementary Figure 1: (A) Example of annotation in QuPath, used to delineate regions of interest which were used for whole transcriptome sequencing in the development and external validation cohorts. (B) Example of annotation in ASAP, applied for spatial transcriptomic samples.
