## Supplementary figures and images for "Predicting bladder cancer molecular subtypes linked to bacillus Calmette-Guerin response from histology images using deep learning"

### Supplementary Figure 2

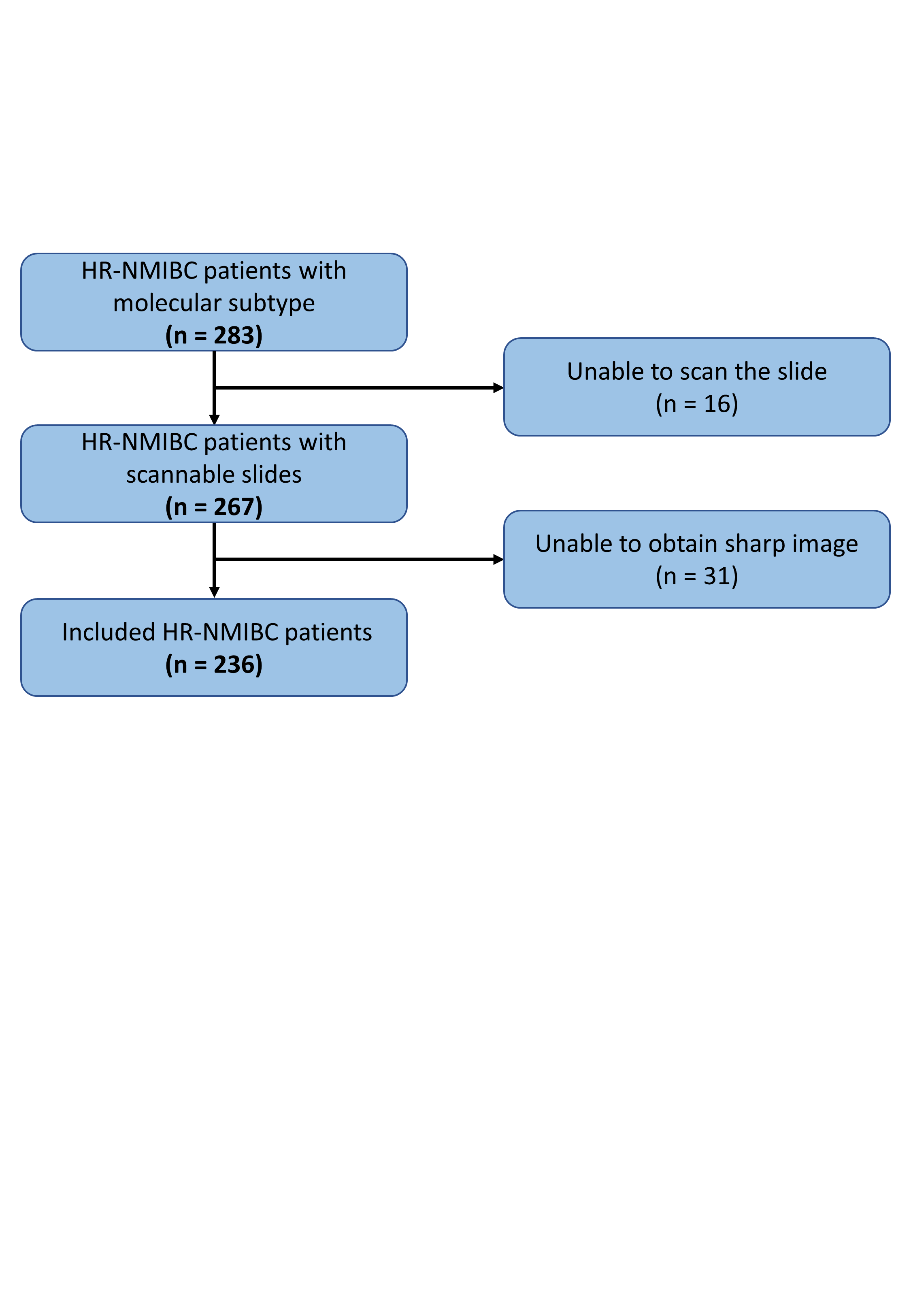

Supplementary Figure 2: The CONSORT chart of the study.
