## Supplementary Figure 3 for "Predicting bladder cancer molecular subtypes linked to bacillus Calmette-Guerin response from histology images using deep learning"

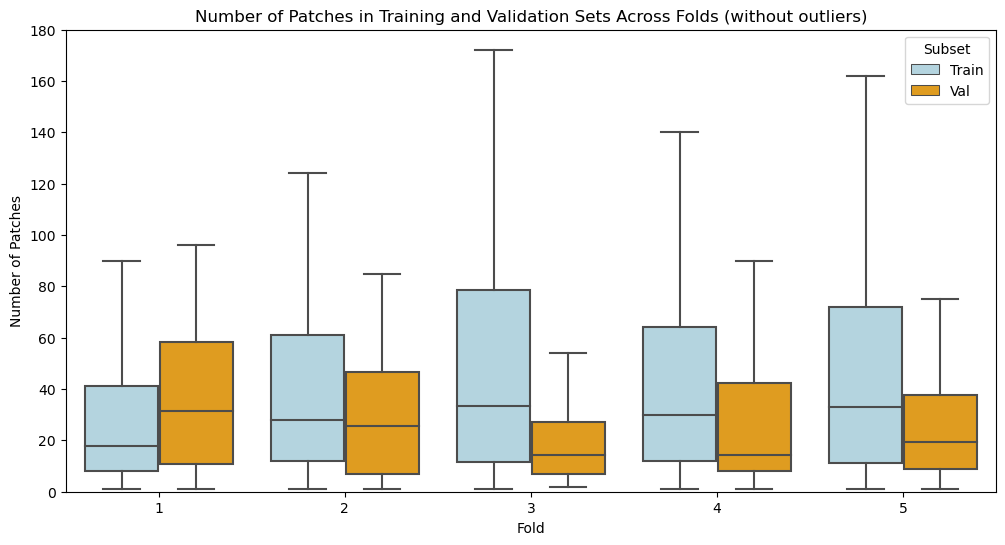

Supplementary Figure 3. Box-and-whisker plots showing the number of image patches per patient in the training and internal-validation sets for each of the five cross-validation folds. Outliers have been removed for clarity. Median, inter-quartile range and full range are indicated; counts are before class oversampling. No significant difference in patch counts per patient was observed between training and validation sets (Kruskal–Wallis test, H = 4.0, p = 0.41).
