## Supplementary Figure 4 for "Predicting bladder cancer molecular subtypes linked to bacillus Calmette-Guerin response from histology images using deep learning"

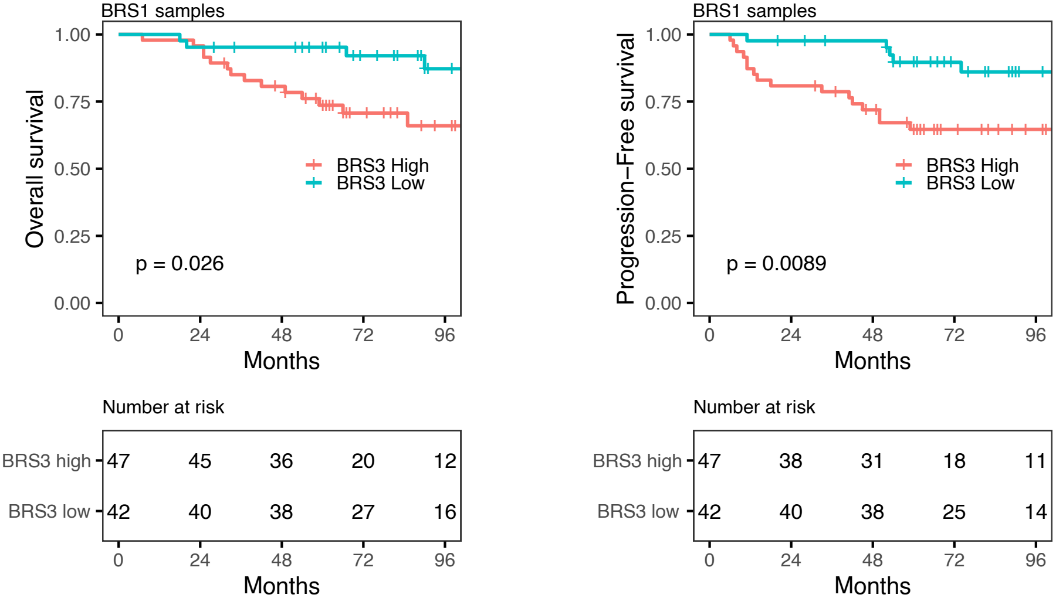
A

B

Supplementary Figure 4: Delineation of intratumor subtype heterogeneity from bulk transcriptomic profiles. (A) Kaplan-Meier plot of progression-free survival (PFS) for 89 patients with BRS2 tumors, stratified by BRS3 weight from WISP analysis (log-rank test). An optimal cutpoint for the BRS fractions according to time to progression was determined using the R package survminer (cutpoint median). (B) Fractions for the four BRS classes for 231 tumors separated by bulk subtype assignment. BRS = BCG response subtype; HR = hazard ratio; CI = confidence interval; RNA-seq = RNA sequencing.

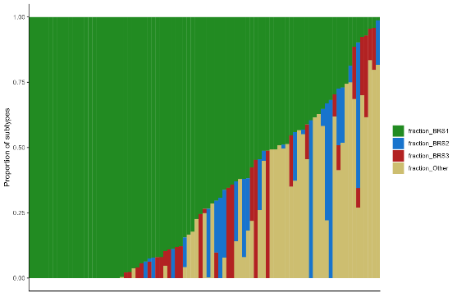

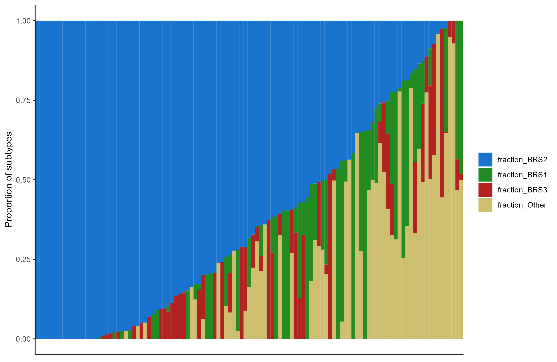

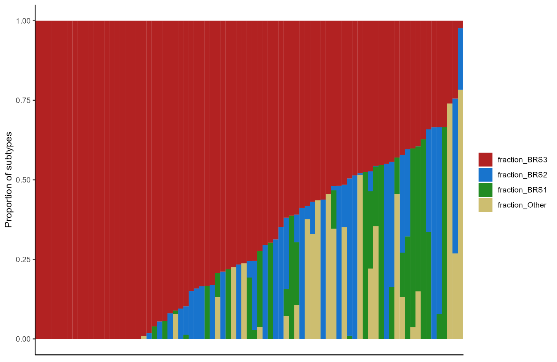

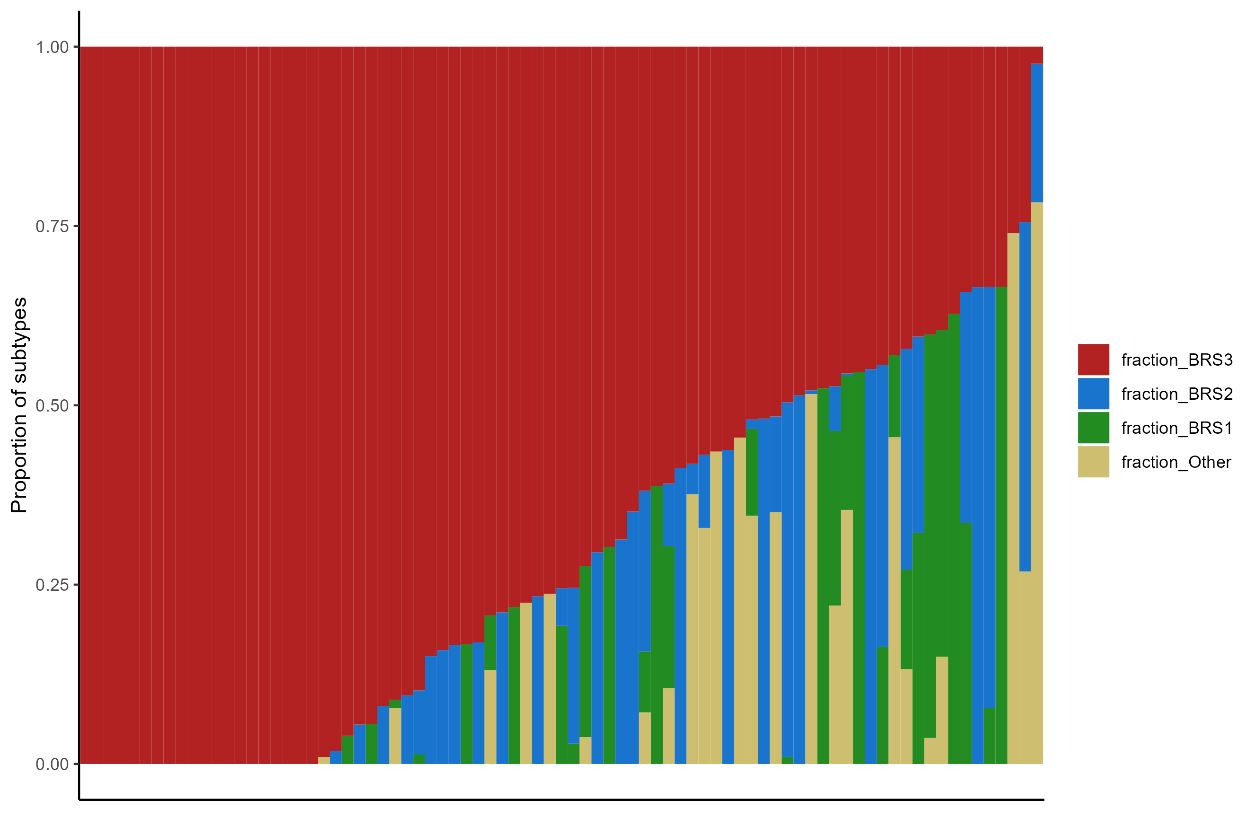
