## Supplementary Figure 5 for "Predicting bladder cancer molecular subtypes linked to bacillus Calmette-Guerin response from histology images using deep learning"

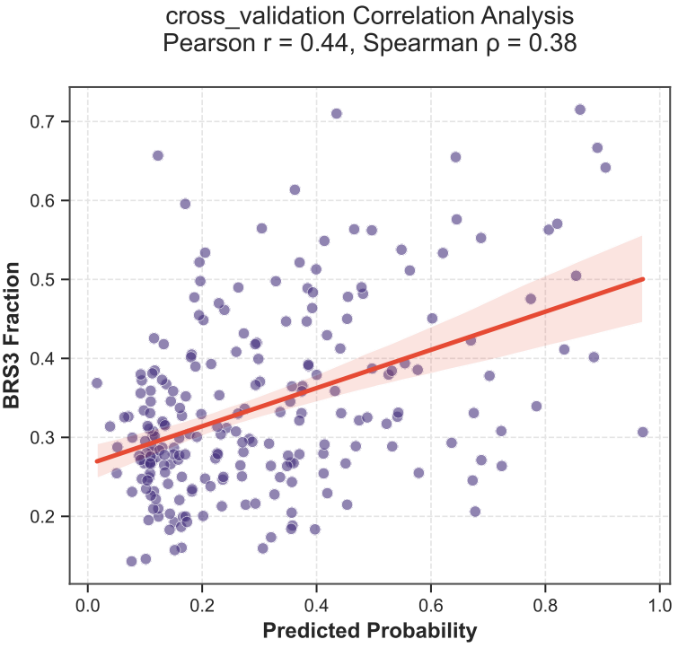

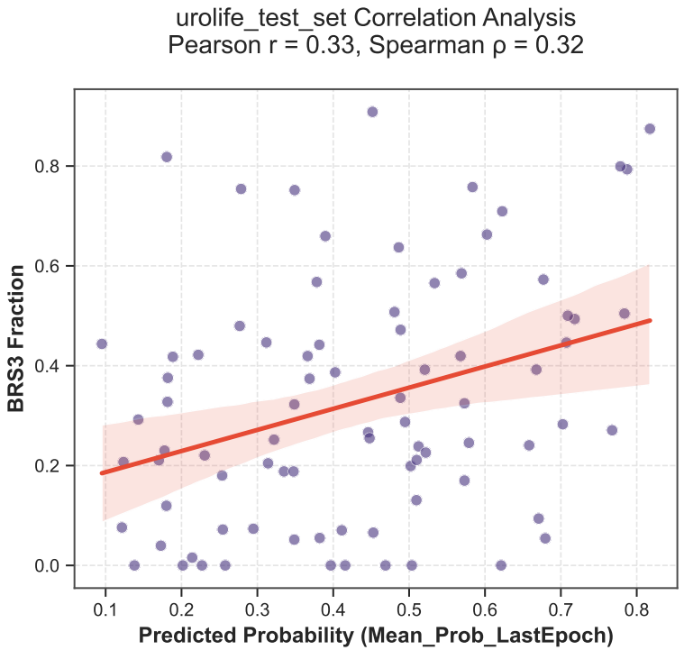
**A	D**

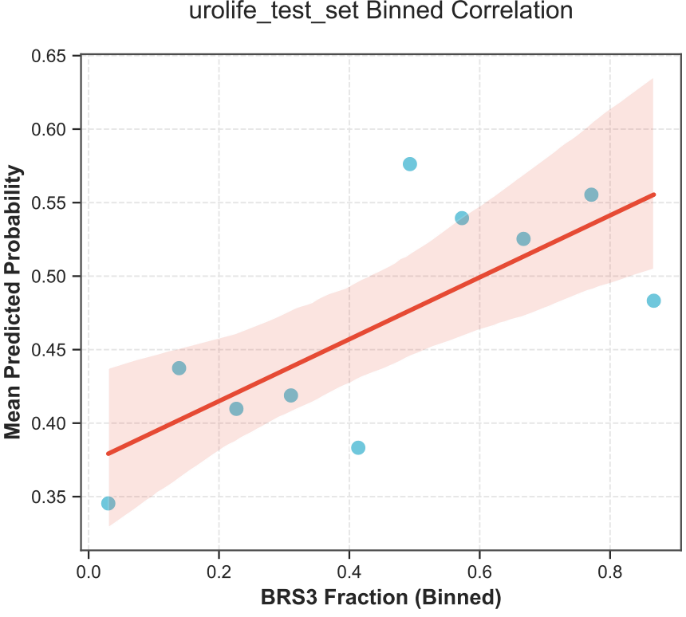

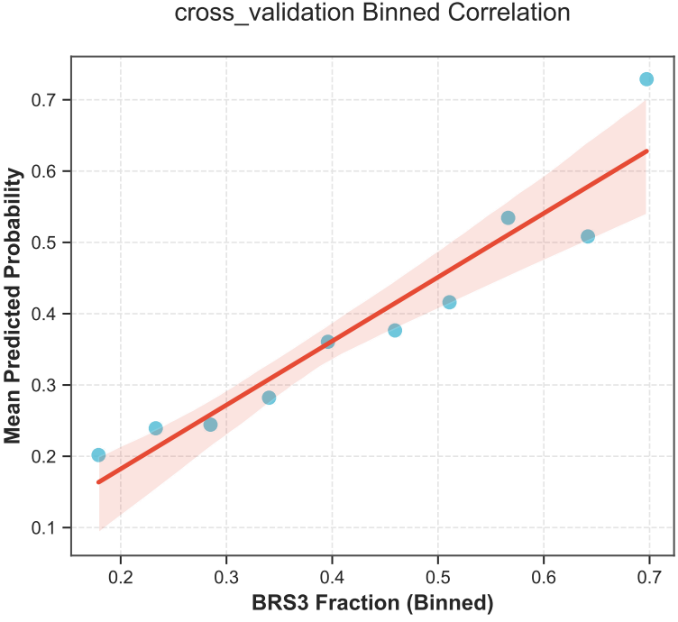
**B	E**

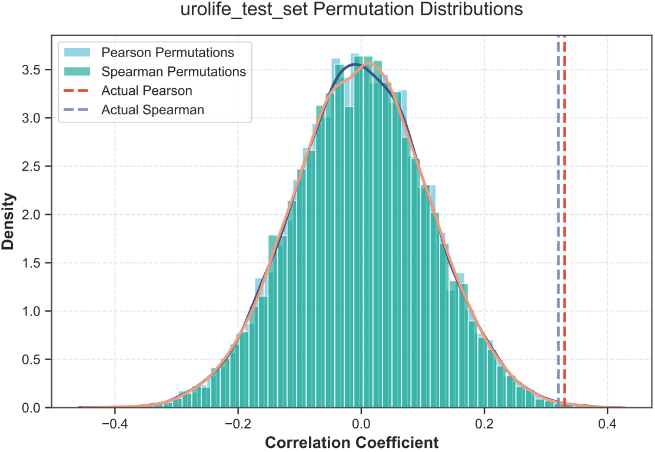

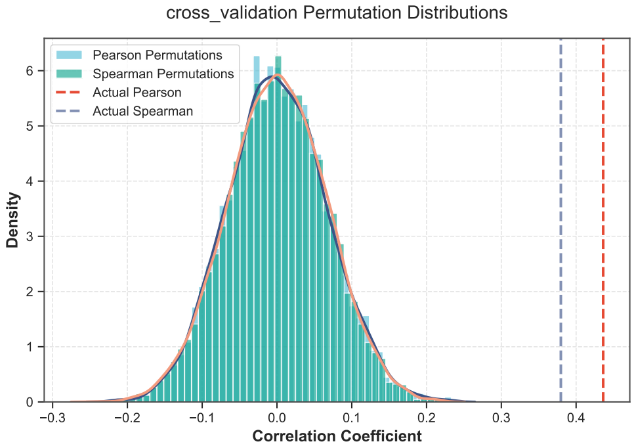
**C	F**

Supplementary Figure 5. Correlation analyses of BMolNet predictions versus true BRS3 fraction: (A–C) and the external validation test set (D–F). (A) Cross‑validation scatter plot. Each point represents one patient’s held‑out prediction (mean probability) plotted against its true BRS3 fraction. The red line is the linear regression fit; Pearson r = 0.44 and Spearman ρ = 0.38 (both p < 0.001). (B) Cross‑validation binned correlation. Patients were sorted by true BRS3 fraction into ten equally sized bins; the blue dots show the mean predicted probability in each bin, and the red line shows the trend across bins, demonstrating that higher BRS3 prevalence corresponds to higher BMolNet scores. (C) Cross‑validation permutation distributions. Histograms display the null distributions of Pearson and Spearman coefficients obtained from 10 000 random shuffles of the BRS3 labels; the dashed red and blue lines mark the observed correlations, which fall in the extreme right tails (empirical p < 0.0001). (D) External validation cohort scatter plot. Held‑out predictions versus measured BRS3 fraction for independent test patients, with regression line (red) and annotated Pearson r = 0.33, Spearman ρ = 0.32 (p = 0.002). (E) External binned correlation. Mean predicted probability is plotted for each of ten BRS3‐fraction bins, illustrating a monotonic increase from the lowest to highest fraction. (F) External permutation distributions. Null distributions of shuffled Pearson and Spearman correlations (10 000 permutations) are shown as histograms; the observed coefficients (dashed lines) lie well beyond the bulk of the permuted values (empirical p = 0.004).
