## Supplementary Figure 6 for "Predicting bladder cancer molecular subtypes linked to bacillus Calmette-Guerin response from histology images using deep learning"

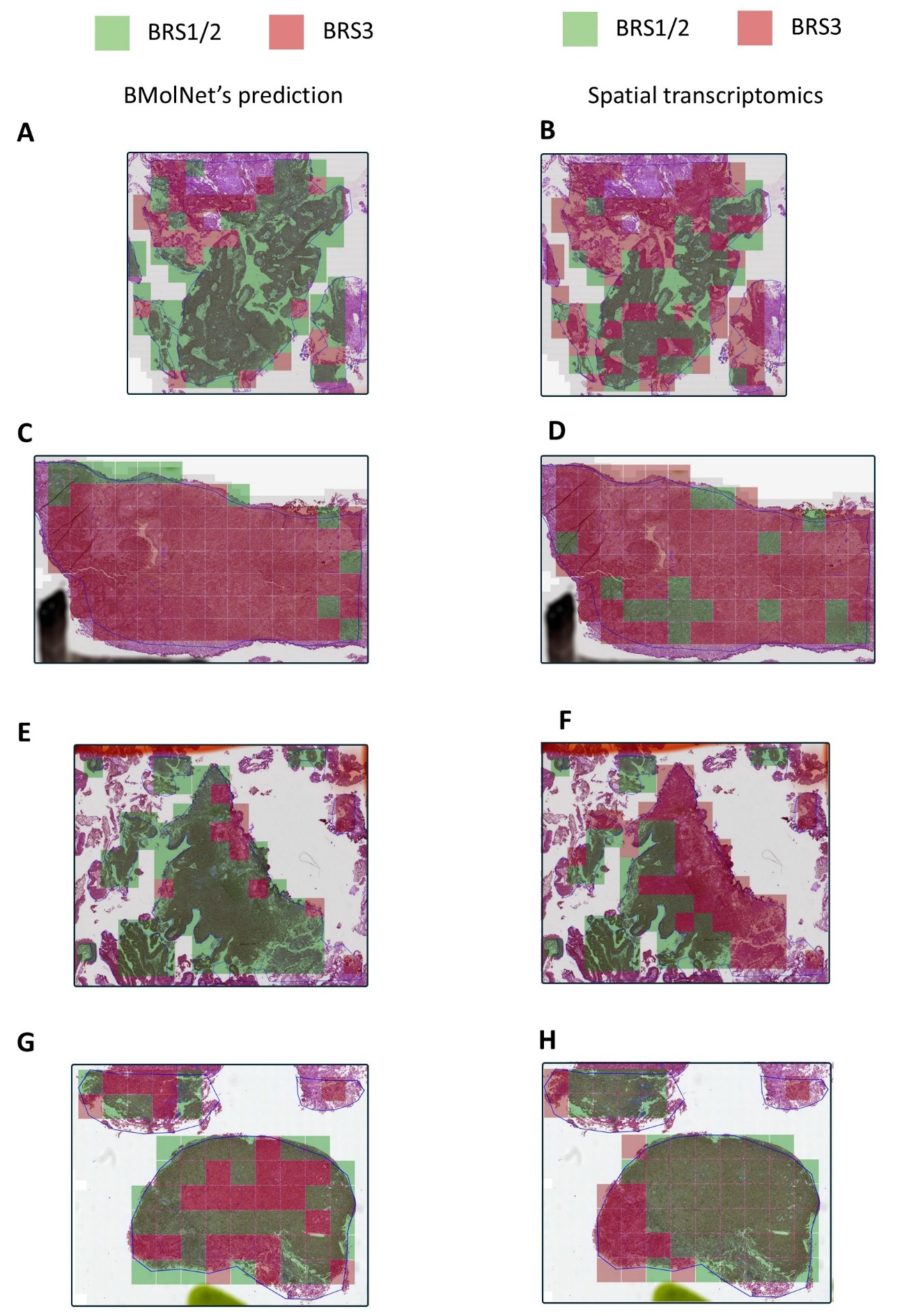

Supplementary Figure 6. Spatial overlays for the four tumors profiled with Visium. Panels A–B, C–D, E–F and G–H correspond to Samples 1, 2, 4 and 5, respectively. Left-hand images in each pair show BMolNet tile-level subtype calls; right-hand images show the transcriptomic BRS labels for the same region.
